# Predicting Imminent Health Outcomes from Common Lab Results

**DOI:** 10.1101/2023.03.01.23286617

**Authors:** Jorge Cerejo, Bernardo Neves, Nuno André da Silva, José Maria Moreira, Mário J. Silva

**Affiliations:** INESC-ID, Instituto Superior Técnico, Universidade de Lisboa, Portugal; Hospital da Luz, Lisboa, Portugal; Hospital da Luz Learning Health, Lisboa, Portugal

**Keywords:** Laboratory Tests, Imminent Hospitalization Prediction, Heart Failure

## Abstract

In recent years, most hospitals have implemented Electronic Health Records to manage and integrate a wide range of medical information, including diagnostics, medication admission and laboratory test results. Certain laboratory variables may serve as indicators of a patient’s clinical deterioration, making laboratory data a valuable tool for identifying high-risk patients. This work introduces a framework for predicting imminent health outcomes (IHO) of multimorbidity patients using laboratory test data. Our cohort includes 322,316 multimorbidity patients that performed laboratory tests in a large teaching hospital between January 2007 and August 2021. Two Imminent Health Outcomes predictive tools were developed. The first considers all patients in the dataset. The second was developed using a subset of patients with Heart Failure (HF) as the main comorbidity (5% of the entire dataset), considering that HF is a highly prevalent syndrome in multimorbidity patients. This predictive model achieved a reasonable predictive performance (*AUROC* = 0.718, 95% CI 0.708-0.756, and *AUPRC* = 0.663, 95% CI 0.630-0.701). C-reactive protein and NT-proBNP are the lab tests that most positively contribute to the prediction of IHO. The IHO predictive tool has the potential to help the medical team identify patients at high-risk of an imminent adverse event, highlighting the laboratory variables that are most contributing to the deterioration of the patient.

## 1 Introduction

Nowadays, patients with two or more chronic conditions, known as multimorbidity patients, are the norm among older people in every healthcare system, mainly due to the increase in life expectancy and population ageing [1]. Furthermore, multimorbidity patients represent 38.3% of the Portuguese population between 24 and 75 years old [2]. Therefore, it is crucial to develop new tools that may help hospitals and physicians understand better these patients’ needs and better plan their medical care. Multimorbidity is highly common in Heart Failure (HF) patients, given that 85% of patients with Heart Failure have concomitant chronic conditions [3].

Electronic Health Records (EHRs) provide extensive medical data with patient medical history. This has motivated an increased interest in developing new technologies and tools in the healthcare sector to improve patient treatment [4]. One example is the prediction of imminent (in less than 24h) health outcomes from common laboratory test results [5].

*Hospital Da Luz Lisboa* (HLL) is a 100% digital hospital that has adopted the use of EHRs since the beginning of its activity. A large amount of health data spanning a fairly significant time-period is available. We had access to a large clinical dataset retrieved from the EHR of HLL under the IntelligentCare (ICare) project, a research initiative of CMU-Portugal [6]. The ICare project aims to create solutions for better treatment of patients with multimorbidities, based on artificial intelligence and advanced analytics methods.

This study aims to research the predictability of imminent health outcomes (IHO) in multimorbidity patients using features extracted from laboratory analysis results. We developed a software tool for predicting imminent hospitalizations of patients admitted to the Emergency Department (ED) room. The tool, named ICIHO (“ICare Imminent Health Outcomes”), can alert which patients are at high-risk and inform better treatment planning.

## 2 Related work

Diagnosis, monitoring, screening, and research are the main goals of using laboratory tests in healthcare [7]. These tests can reduce diagnostic errors, adding value to decision-making in healthcare [8]. Laboratory tests may be used as an indicator of adverse events, since abnormal values of laboratory tests are associated with causes of mortality and morbidity. Also, it has been shown that laboratory results can predict IHO of patients admitted to the ED [5].

The need for health data exchange between clinical information systems has motivated the development of standards for processing lab tests data. Logical Observation Identifiers, Names and Codes (LOINC) is the international common language used for the identification and representation of laboratory measurements, facilitating data exchange and interoperability [9]. The Observational Medical Outcomes Partnership (OMOP) Common Data Model (CDM) is an informational model that enables the conversion of health data from several sources, namely EHRs, with pre-specified and standardized coding schemes, terminologies and vocabularies. Therefore, clinical studies based on EHRs data are better supported by OMOP CDM given the uniformization of the health data [10].

In the last few years, there has been a growing body of literature regarding the prediction of IHO from laboratory results. Machine learning (ML) models, primarily Logistic regression (LR), are used in this analysis. More recently, some novel studies use models based on deep learning, in which neural networks (NN) have been trained to predict outcomes [11].

Loekito et al. (2013) constructed a multivariate LR model to predict IHO. Amongst 30 laboratory variables, they selected the 9 most statistically significant variables. To assess the performance of the model, they computed the area under the receiver operating characteristic curve (*AUROC*). The model showed a reasonable (*AUROC* = 0.69), good (*AUROC* = 0.82) and very good (*AUROC* = 0.90) predictive ability for the prediction of imminent Medical Emergency Teams (MET) calls, Intensive Care Unit (ICU) admission and death, respectively [5].

Mueller et al. (2020) developed four different ML models to predict in-hospital mortality. The second model is the most significant since includes laboratory data and achieved a good performance (*AUROC* = 0.88). Moreover, a model with demographic and laboratory data was trained to predict hospitalizations, showing a good predictive ability (*AUROC* = 0.80) [12].

Seki et al. (2021) developed an ML-based model to predict in-hospital death in a short-term period (14 days). The researchers constructed an NN, in which demographic (age and gender) data and 21 laboratory variables were used. The authors concluded that it was possible to predict in-hospital mortality with high predictive performance (*AUROC* = 0.942). Since the dataset was imbalanced, they also computed the area under the precision-recall curve (*AUPRC*), obtaining an *AUPRC* = 0.191 [13].

## 3 Data Processing

Figure 1 provides the visual overview of the workflow adopted in this study, including the development of the ICIHO predictive tool, which will be discussed in Section 4.

**Fig. 1:**
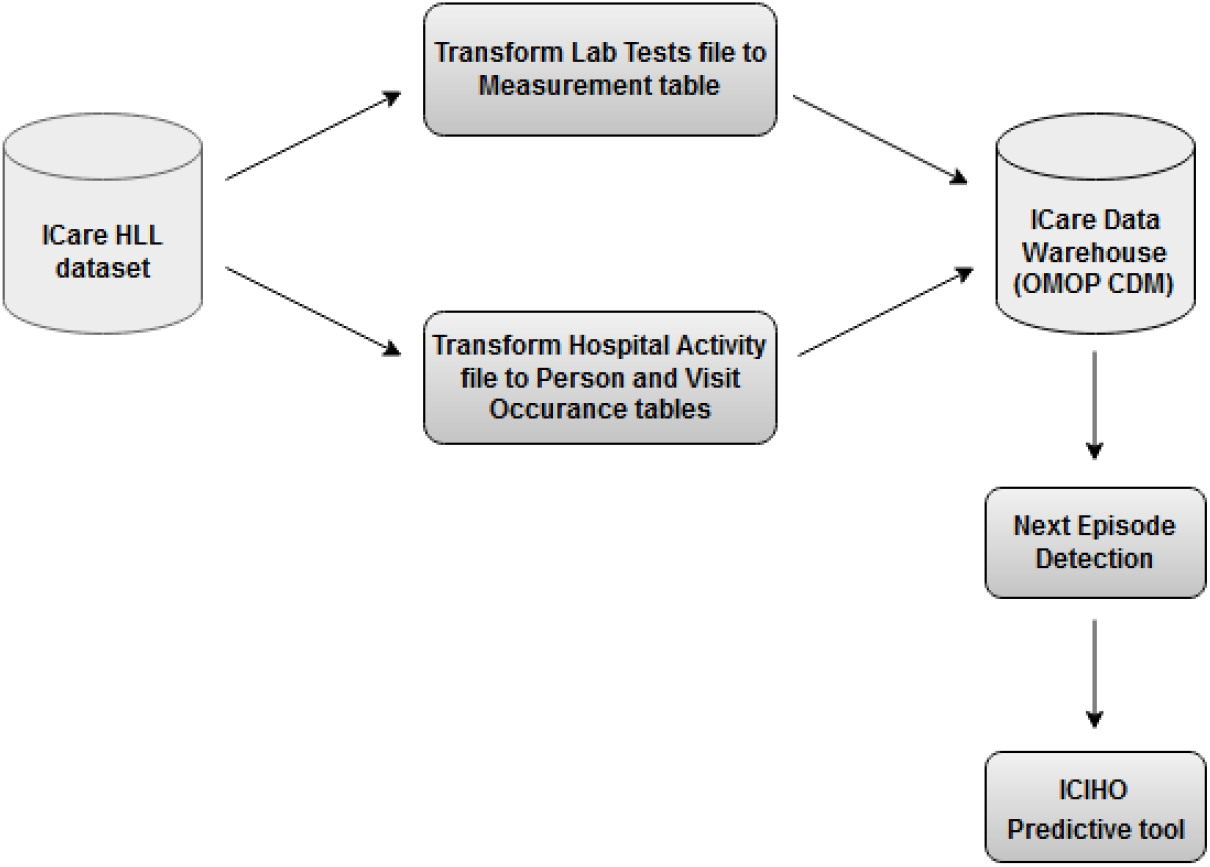
Overview of the workflow adopted in this study.

The IntelligentCare (ICare) dataset is a collection of (csv) files containing data extracted from EHR of the HLL. The ICare dataset contains 26 files with data on the medical histories of 834,529 patients and an observation period spanning between January 2007 and August 2021.

Our data pre-processing work focused on the laboratory tests file, which contains several fields including laboratory tests and their results, along with the ID of the patient and the episode in which the laboratory test was ordered (Table 1). As not all patients performed at least one laboratory test in the observation period, our cohort includes 322,316 patients (39% of the ICare dataset patients).

**Table 1:** Fields of laboratory test table and their description.

| Table Field | Description |
| --- | --- |
| NHC ("*Número Histórico Clínico*") | Code that identifies the patient's clinical history |
| Episode Identifier | Code that uniquely identifies the episode |
| Date Result | Date and time the laboratory test was performed |
| Test Code | Code that identifies the laboratory test |
| Test Description | Description of the laboratory test |
| Reference Value | Interval in which the laboratory result is normal |
| Unit | Unit in which the laboratory test is measured |

### 3.1 Lab Test Codes Harmonization

The lab test codes in the ICare Dataset are not uniformly coded across the (long) period of observation. Despite being LOINC-based, they do not always directly match. Hence, a necessary first step in preparing the dataset was mapping every test name to the LOINC international standard. The LOINC mapping is implemented as a simple script developed to convert local HLL laboratory codes to LOINC codes.

We implement indirect mapping strategies, when a HLL code needs some tweaking (see Table 2).

**Table 2:** Examples of the different mapping types that exist between HLL laboratory codes and LOINC codes.

| Type of Mapping | HLL lab code | LOINC code |
| --- | --- | --- |
| Direct mapping | 5770-3 | 5770-3 |
| Indirect mapping | 789-8-4 | 789-8 |
| No mapping | 236-17003 | --- |

The mapping script takes as input the HLL laboratory codes and the LOINC standardized codes table. Then, the script applies various heuristics, identifying the mapping strategy to be applied (for indirect mapping, the last digit is removed). Finally, it returns a table with the HLL laboratory and LOINC codes.

The tests in the ICare dataset which we cannot map to a LOINC code correspondence are excluded from the dataset.

The mapping between HLL laboratory codes and LOINC codes was successfully performed for 2041 codes, covering 88.60% of the laboratory tests dataset. 1307 codes (63.78% of the dataset) were mapped directly, whereas 734 codes (24.82% of the dataset) were obtained via indirect mapping. We identified 1510 HLL laboratory codes for which we could not assign a corresponding LOINC code, covering 11.40% of the laboratory tests dataset.

### 3.2 Laboratory Results Processing

The laboratory tests dataset includes multiple types of results, some of them manually introduced in free text fields in the EHR.

The results of laboratory tests are mostly quantitative or nominal. A processing step was also developed to clean and harmonize the values of laboratory tests results. Therefore, we transformed the data so that quantitative results can be interpreted as numerical values and that categorical results with the same meaning can be represented by a unique nominal label.

The results of the laboratory tests were harmonized using regular expressions. After this step, the laboratory values that were not harmonized (7.11% of the laboratory test results) were discarded.

Of the laboratory test data that was not discarded, 36.69% of laboratory results did not require any modification. By contrast, 63.31% of laboratory results needed to be transformed.

### 3.3 Loading Measurements Data into the ICare DataWarehouse

The ICare project has created an OMOP CDM (version 5.3) Datawarehouse [14]. We populated the OMOP CDM *Measurement* table of the ICare Datawarehouse with the processed laboratory tests data from the Icare dataset. The *Measurement* table data has references to information on the patients (table *Person*), and hospital activity data (table *Visit Occurrence*).

The *Measurement* table includes 32,917,962 measurements, regarding laboratory tests performed by 302,970 patients in 1,045,327 episodes. Of these episodes, 667,825 are consultations, 310,285 are Emergency Department (ED) admissions and 67,217 episodes correspond to hospitalizations.

### 3.4 Next Episode Detection

To predict IHO using laboratory results, it is necessary to link the laboratory measurements to the next episode in each patient history.

We developed a simple script with an algorithm for identifying the clinical episode that follows the laboratory measurements of a patient. The Next Episode Detection algorithm receives the Measurement and the Visit Occurrence tables from the ICare database and, based on the time stamps and the episode of each laboratory measurement, returns the next episode and the time difference between them.

## 4 Imminent Health Outcomes Prediction

This section provides a description of the proposed workflow for the development and evaluation of the ICIHO predictive tool, using the HLL processed data in the ICare Datawarehouse. We predict imminent (*<*24 hours) hospitalizations of patients admitted to the ED. In Section 4.1, we use all patients from the ICare dataset. In contrast, Section 4.2 is disease-specific, restricting the scope of the predictive model to multimorbidity patients with heart failure as the main comorbidity.

### 4.1 ICIHO as Predictive Tool

There are 310,285 ED admissions in the ICare dataset, of which 33,084 (10.7%) are followed by an imminent hospitalization (i.e., the next episode is a hospitalization in less than 24 hours). These episosdes are labelled as positive (“1”). Contrary, in 277,201 episodes, patients were discharged from ED and the episode is therefore labelled as negative (“0”).

The measurement table of the ICare Data Warehouse includes 1002 different laboratory tests. Considering the prescription/non-prescription of laboratory tests, a chi-square test showed that 227 laboratory variables are statistically significant in predicting imminent hospitalizations. Univariate LR revealed that only 70 variables (69 laboratory tests and age) are statistically significant when the laboratory test results are considered. The adjusted significance level after Bonferroni correction is *α* = 0.0001.

Nevertheless, combining these variables with the ones used in [5] and selecting those that represent at least 10% of the laboratory measurements, 20 features were selected to be included in the ICIHO predictive model. Hemoglobin (LOINC code 718-7) is highly correlated with Hematocrit (LOINC code 4544-3), hence, it was not used in the model training.

#### Multivariate LR model based on the laboratory tests prescription

Before predicting IHO from common laboratory results, a Multivariate LR model was constructed based on the prescription/non-prescription of 227 laboratory tests. Table 3 presents the performance of this model.

**Table 3:** Performance metrics of the multivariate LR model trained based on the laboratory tests prescription.

| Metric | Value [95% CI] |
| --- | --- |
| Balanced accuracy | 0.756 [0.751 - 0.762] |
| Precision | 0.288 [0.281 - 0.295] |
| Recall | 0.727 [0.716 - 0.738] |
| F1-score | 0.412 [0.404 - 0.420] |
| AUROC | 0.825 [0.820 - 0.831] |
| AUPRC | 0.463 [0.451 - 0.475] |

This Multivariate LR model has a low precision (0.288). However, it is worth pointing out the high recall value of this classifier (0.727) since the well-being of patients is at risk. Moreover, the IHO model presents good values of balanced accuracy and AUROC. The last metric may be biased by the imbalanced dataset and it is not reliable for this case. Considering the AUPRC, the model performance is poor (0.463).

Despite its low performance, this tool can be important in a clinical context to help physicians flag early on which patients are most at risk of hospitalization, as it does not require laboratory test results.

#### IHO prediction from common laboratory results

Previously, LR was employed to investigate the potential influence of laboratory test prescription (or non-prescription) on the prediction of IHO. In this task, we compared LR and NN to predict IHO from common laboratory results. The model choice is a trade-off between complexity (and probably a better performance by the neural network) and explainability (given that LR coefficients are easier to interpret).

Bearing in mind that real clinical data is being used, the dataset presents some problems, such as imbalanced classes and missing values. To deal with these problems, different approaches are used during the training of the model to optimize its performance.

The first problem relies on the trade-off between the number of features and the number of samples. This work considered three prescription proportion values, namely 10%, 40% and 60%.

Undersampling and class weight correction are the approaches used to deal with the problem of imbalanced classes. Regarding the missing values, two approaches were considered, namely imputation and deletion of episodes with null values. Imputation was performed based on Multivariate Imputation Chain Equations (MICE) technique [15].

To avoid overfitting, Elastic Net regularization penalty was used during the LR-based model training. The parameters were optimized using 5-fold cross-validation, in which f1-score was the metric chosen to optimize the model performance (since it combines precision and recall). The neural network architecture consists of two hidden layers, each with four neurons, following the design proposed by Dervishi [16]. The loss function used is the cross-entropy and the model parameters were optimized using the stochastic gradient descendent. The weights were updated using backpropagation algorithm.

The different models were evaluated using common performance metrics: precision, recall, f1-score, balanced accuracy, AUROC and AUPRC. Table 5 summarizes the performances of the different approaches considered in this section. As shown, all approaches perform similarly, with NN slightly outperforming LR-based models. Nonetheless, NN are more complex than LR, namely NN requires optimizing 2 *∗* 4 *∗ N features* (2 hidden layers and 4 neurons per layer) parameters, while the number of parameters used by LR is equal to the number of features.

Moreover, models with a proportion threshold of 10% achieved the best performance metrics. However, due to the high number of features, only a few records were available. Thus, this model may not generalize well for other datasets. A proportion threshold of 60% represents a model trained only with seven features, which can explain the decrease in the model performance. Therefore, we considered the proportion threshold of 40% a good trade-off between the number of features and episodes available.

Furthermore, class weight correction outperforms undersampling, adding the advantage that it does not exclude negative samples. Therefore, we considered class weight correction a better strategy for dealing with the imbalanced class problem.

The proportion of negative samples increases when the imputation strategy is adopted, affecting the model’s performance, as shown by the values of the f1-score and AUPRC columns (Table 5). Therefore, strategies without imputation are better approaches and the models trained present reasonable performances in predicting imminent hospitalizations.

Figure 2 compares the performances of both models (LR and NN) when the proportion threshold equals to 40%, class weight correction is adopted and missing values are not imputed.

**Fig. 2:**
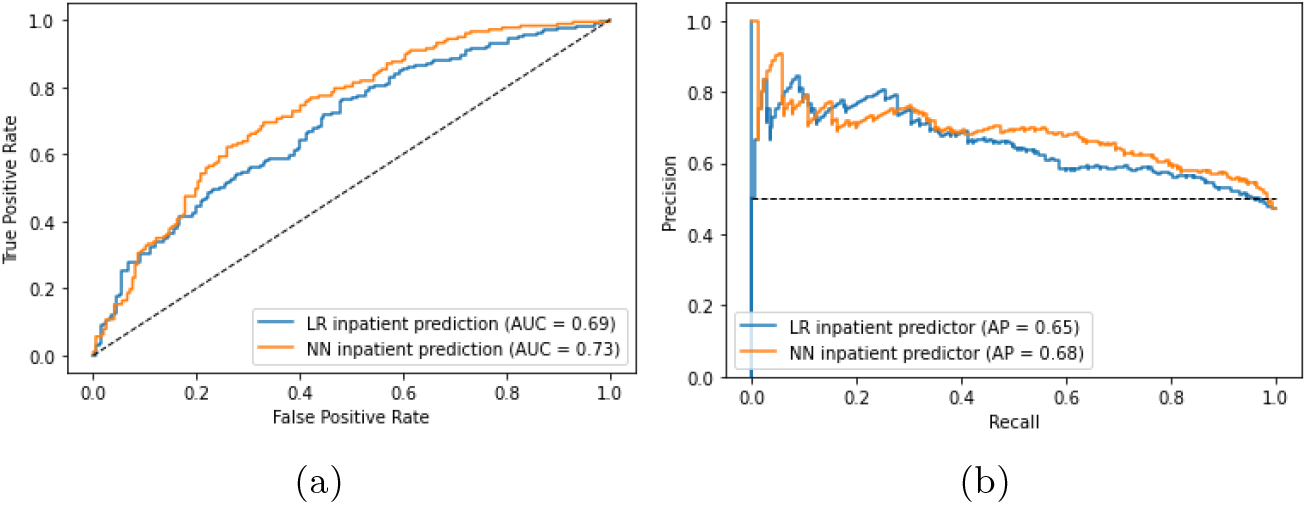
Plot of (a) ROC and (b) Precision-recall curves to compare the performances of LR-based model with the NN-based model for the prediction of IHO based on the laboratory tests results.

The models reported in the literature performed slightly better than the ones presented here. Nevertheless, those models only present AUROC curve values to assess model’s performance, which in our study is not always a reliable metric [5], [12].

### 4.2 ICIHO as Heart Failure Predictive Tool

We specifically analyzed a subset of the total population, by including only patients with HF and concomitant comorbidities. This cohort was selected based on ICD-9 codes used to identify HF syndrome that were defined in a previous study within the ICare group [14]. There are 3,908 HF patients identified in the ICare dataset, amongst which 3,407 have at least one laboratory test performed and available on the dataset. The *Measurement* table includes 2,181,744 laboratory tests performed (about 5.5% of the entire dataset). There are 657 different laboratory tests performed on HF patients.

There are 46,922 different episodes in which laboratory tests were performed. These episodes account for 27,744 outpatient appointments, 12,689 ED admissions and 6488 hospitalizations. Considering the ED episodes, 4693 (37.0%) are followed by a hospital admission in less than 24 hours.

The workflow for the construction of the HF-ICIHO predictive tool is identical to ICIHO presented in Section 4.1. Due to the lower number of samples, the proportion thresholds were modified for 50% (19 features), 75% (16 features) and 90% (9 features). Class weight correction was adopted and NT-proBNP was the only imputed laboratory variable (median value imputation), given the importance of this biomarker in the prevention, assessment and management of HF.

Table 6 summarises the performances of the six models trained for constructing the HF-ICIHO predictive tool. Figure 3 shows the comparison between the models considering the ROC (a) and precision-recall (b) curves.

**Fig. 3:**
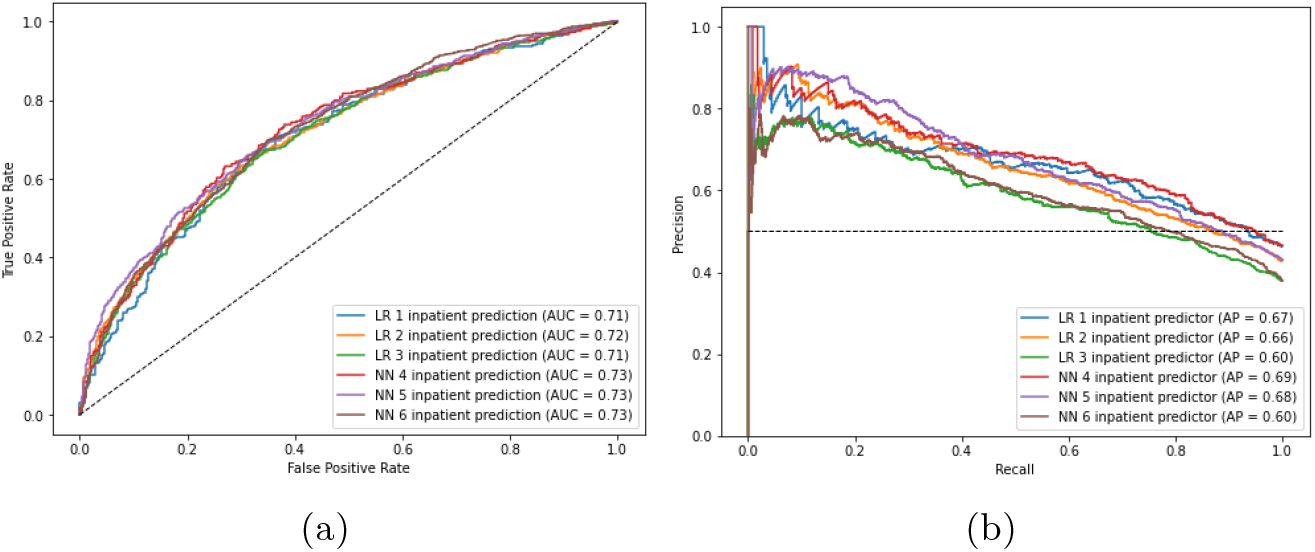
Comparison of (a) ROC and (b) PR curves for the models trained.

The performances of the several models are similar, and overall the NN slightly outperforms the LR. Models using more features (HF1 and HF4) have a better performance. However, these models include fewer samples. Models with a small number of features (HF3 and HF6) show a decrease in performance metrics.

Considering the trade-off complexity vs model performance, we believe that the HF2 model (highlighted in Table 6) provides the best approach for implementing the HF-ICIHO predictive tool, given the simplicity of the LR. This model presents a reasonable f1-score (0.617), as well as AUROC (0.718) and AUPRC (0.663) values. Even though the recall (0.624) value is not poor, a higher recall would be desirable given the clinical context in which this study is inserted.

The coefficients of the LR model can be interpreted by computing the odds ratio, which can be calculated by exponentiating the coefficients of the model [17]. An odds ratio greater than 1 indicates an increased likelihood of imminent hospitalization (i.e., positive class), whereas a value less than 1 contributes to the prediction of the negative class (i.e., not imminent hospitalization).

Therefore, Table 4 presents the odds ratio of the coefficients for the HF2 model. The results show that high values of C-reactive protein and NT-proBNP significantly contribute to positive outcomes, i.e., the prediction of imminent hospitalization for patients admitted to the ED. Conversely, low values of Lymphocytes and Erythrocytes negatively contribute to the prediction of imminent hospitalization.

**Table 4:**
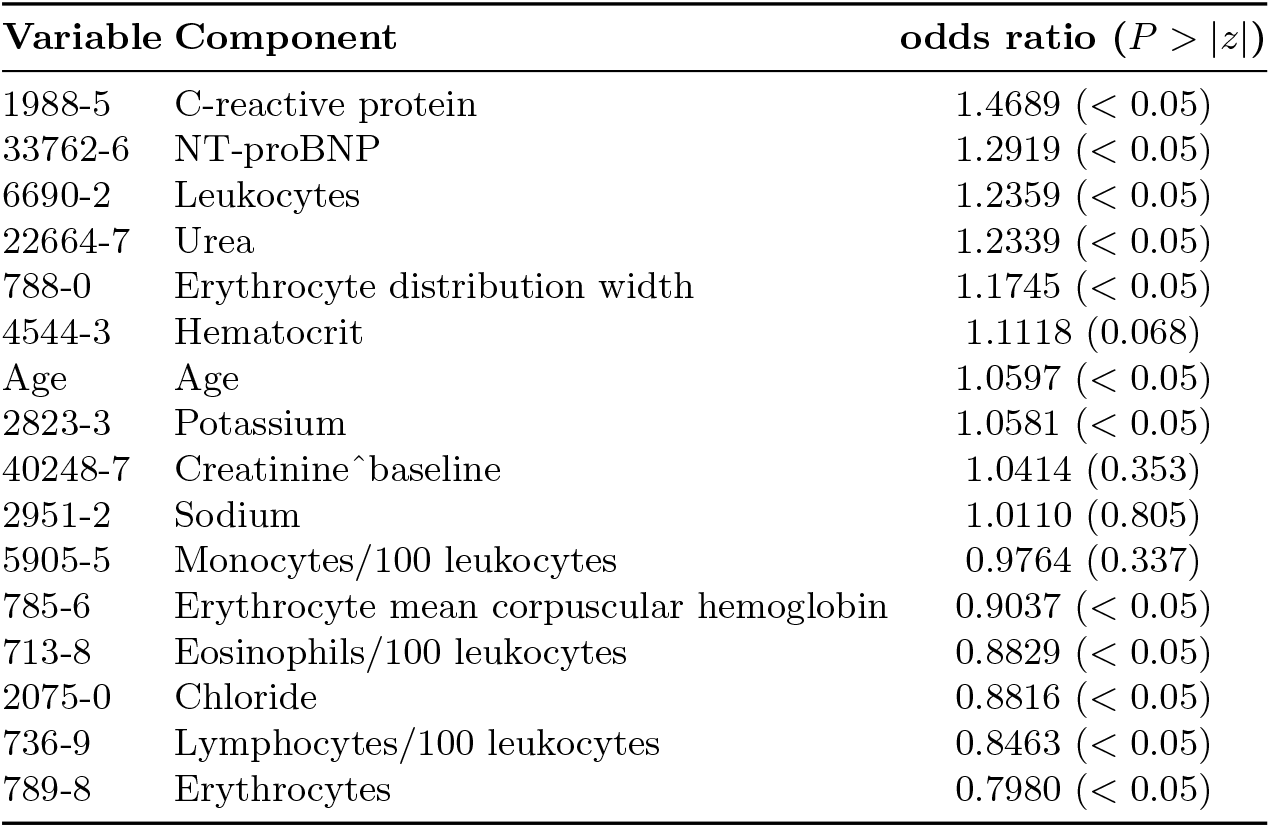
Interpretation of Model’s Coefficients based on the odds ratio of each coefficient. Wald test was performed to compute the p-values for the null hypothesis that the coefficient is equal to zero, i.e., the odds ratio is equal to one.

**Table 5:**
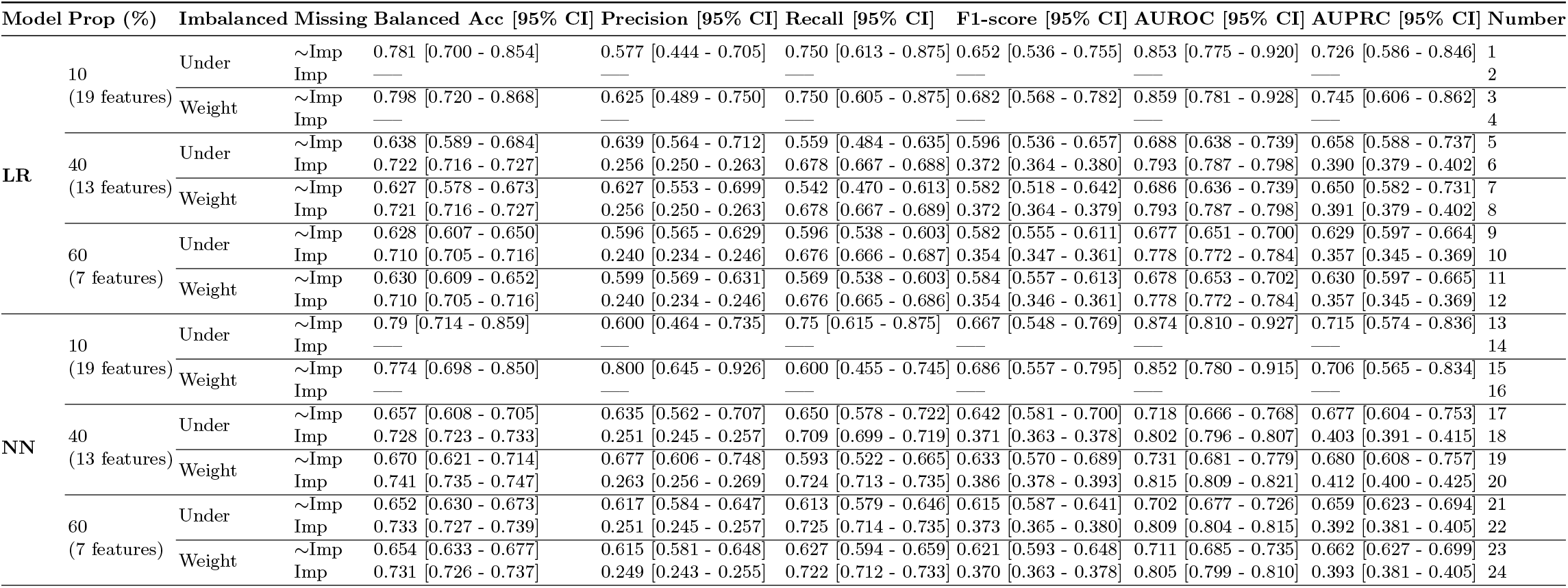
Summary of the performance metrics used to assess the models trained for different approaches using the entire dataset.

**Table 6:**
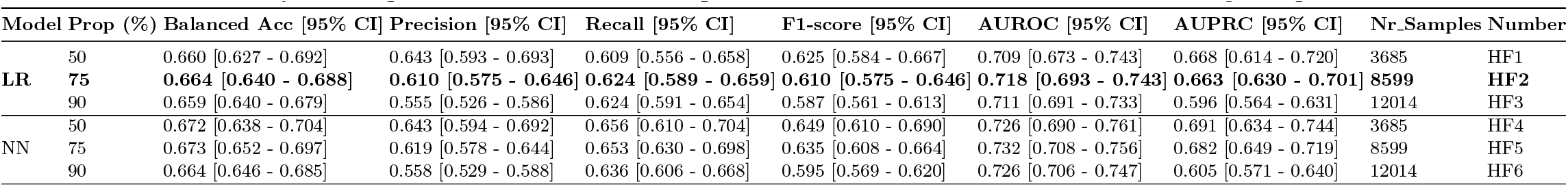
Summary of the performance metrics computed to evaluate the models trained using HF patients dataset.

Table 4 displays the p-values associated with each coefficient of the multivariate model. Smaller p-values indicate stronger evidence against the null hypothesis, suggesting that the corresponding coefficients are more relevant to the outcome (imminent hospitalization). Variables such as Hematocrit, Creatinine, Sodium, and Monocytes were found to be insignificant to the outcome.

## 5 Conclusions

We proposed a framework to predict imminent health outcomes for patients with multimorbidity. Using data processing techniques and machine learning algorithms applied to laboratory test data from *Hospital da Luz Lisboa*, we were able to stratify patients at higher risk of adverse events.

This retrospective study led to the identification of the most predictive laboratory tests in predicting a hospitalization in the following 24 hours, when presenting in the ED, therefore helping the medical team understand which patients are more at risk, and allowing for better resource allocation.

This analysis is robust, considering the different approaches used to study the best strategy for ICIHO implementation. Nevertheless, it would benefit from an external validation with datasets from other hospitals.

This study also emphasizes the importance of explainability in machine learning models. As observed, the LR provides a straightforward interpretation of the weight of each feature. On the other hand, NN do not provide an easily interpretable understanding of features weights, due to their complexity and black-box nature. Furthermore, the results indicated that the performance of NN was not significantly superior when compared to LR. Given this, the preference for a more explainable model was deemed to be a more favorable trade-off.

A limitation of this work is related to the restricted use of laboratory variables. That is, we believe that other medical data (vital signs, diagnoses, symptoms, other exams such as radiology) may increase the performance of the models trained. We intend to to use this data in future works to confirm this hypothesis.

Another limitation relies on the format of the input data, that is, only the laboratory data of the actual episode is used to predict the next one. However, we are convinced that data about previous episodes may also be significant. For that, the use of Long Short-Term Memory (LSTM) networks should be considered.

Despite these limitations, this study provides evidence that laboratory values are useful for risk prediction of hospital admission. The developed framework can therefore be adapted for different event predictions or different time windows, as well as for predicting IHO in other populations, such as patients with Diabetes, Chronic Kidney Disease (CKD) and Chronic Obstructive Pulmonary Disease (COPD).

## Data Availability

All data produced in the present study are available upon reasonable request to the authors

## Acknowledgments

This work was developed by the IntelligentCare project LISBOA-01-0247-FEDER-045948 that is co-financed by the ERDF/LISBOA2020 and by FCT under CMU-Portugal and by FCT under projects UIDB/50021/2020, UIDB/00408/2020, UIDP/00408/2020.

We thank Rui Lopes Baeta for the assistance in loading the lab test data into the ICare Data Warehouse. Predicting Imminent Health Outcomes from Common Lab Results 13

## References

[1] WHO, “Multimorbidity Technical Series on Safer Primary Care Multimorbidity: Technical Series on Safer Primary Care,” p. 28, 2016. [Online]. Available: http://apps.who.int/bookorders.

[2] G. Q. Romana, I. Kislaya, M. R. Salvador, S. Cunha-Goncalves, B. Nunes, and C. Dias, “Multimorbidity in portugal: Results from the first national health examination survey,” Acta Médica Portuguesa, vol. 32, no. 1, 2019. DOI: 10.20344/amp.11227. [Online]. Available: https://www.actamedicaportuguesa.com/revista/index.php/amp/article/view/11227.

[3] A. M. Chamberlain, J. L. Sauver, Y. Gerber, et al., “Multimorbidity in Heart Failure: A Community Perspective,” The American journal of medicine, vol. 128, no. 1, p. 38, 2015, ISSN: 15557162. DOI: 10.1016/J.AMJMED.2014.08.024. [Online]. Available: https://www.amjmed.com/article/S0002-9343(14)00790-6/fulltext.

[4] S. Dash, S. K. Shakyawar, M. Sharma, and S. Kaushik, “Big data in health-care: management, analysis and future prospects,” Journal of Big Data, vol. 6, no. 1, pp. 1–25, Dec. 2019, ISSN: 21961115. DOI: 10.1186/S40537-019-0217-0/FIGURES/6. [Online]. Available: https://journalofbigdata.springeropen.com/articles/10.1186/s40537-019-0217-0.

[5] E. Loekito, J. Bailey, R. Bellomo, et al., “Common laboratory tests predict imminent medical emergency team calls, intensive care unit admission or death in emergency department patients,” Emergency Medicine Australasia, vol. 25, no. 2, pp. 132–139, 2013. DOI: 10.1111/1742-6723.12040. [Online]. Available: https://onlinelibrary.wiley.com/doi/abs/10.1111/1742-6723.12040.

[6] Intelligentcare, Oct. 2022. [Online]. Available: https://www.cmuportugal.org/large-scale-collaborative-research-projects/intelligentcare/.

[7] F. H. Wians, “Clinical laboratory tests: Which, why, and what do the results mean?” Laboratory Medicine, vol. 40, no. 2, pp. 105–113, 2009. DOI: 10.1309/LM4O4L0HHUTWWUDD. [Online]. Available: https://academic.oup.com/labmed/article/40/2/105/2504825.

[8] M. Plebani and G. Lippi, “Improving diagnosis and reducing diagnostic errors: The next frontier of laboratory medicine,” Clinical Chemistry and Laboratory Medicine, vol. 54, no. 7, pp. 1117–1118, 2016. DOI: 10.1515/cclm-2016-0217. [Online]. Available: https://www.degruyter.com/document/doi/10.1515/cclm-2016-0217/html.

[9] Loinc® Indianapolis, IN: Regenstrief Institute, Inc., Logical Observation Identifiers Names and Codes (LOINC). [Online]. Available: https://loinc.org/.

[10] B. E. Dixon, C. Wen, T. French, J. L. Williams, J. D. Duke, and S. J. Grannis, “Observational Health Data Science and Informatics (OHDSI),” BMJ Health Care Inform, vol. 27, p. 100 054, 2020. [Online]. Available: https://ohdsi.github.io/TheBookOfOhdsi/CommonDataModel.html#fn18.

[11] F. Shamout, T. Zhu, and D. Clifton, “Machine learning for clinical outcome prediction,” IEEE Reviews in Biomedical Engineering, vol. PP, pp. 1–1, Jul. 2020. DOI: 10.1109/RBME.2020.3007816.

[12] O. Mueller, K. Rentsch, C. Nickel, and R. Bingisser, “Disposition decision support by laboratory based outcome prediction,” Journal of Clinical Medicine, vol. 10, p. 939, Mar. 2021. DOI: 10.3390/jcm10050939. [Online]. Available: https://www.mdpi.com/2077-0383/10/5/939.

[13] T. Seki, Y. Kawazoe, and K. Ohe, “Machine learning-based prediction of in-hospital mortality using admission laboratory data: A retrospective, single-site study using electronic health record data,” PLOS ONE, vol. 16, no. 2, e0246640, Feb. 2021, ISSN: 1932-6203. DOI: 10.1371/JOURNAL.PONE.0246640. [Online]. Available: https://journals.plos.org/plosone/article?id=10.1371/journal.pone.0246640.

[14] R. Baeta, “Identifying clinical trajectories in patients with multimorbidity,” M.S. thesis, Instituto Superior Técnico, Oct. 2022.

[15] M. J. Azur, E. A. Stuart, C. Frangakis, and P. J. Leaf, “Multiple imputation by chained equations: what is it and how does it work?” International Journal of Methods in Psychiatric Research, vol. 20, no. 1, p. 40, Mar. 2011, ISSN: 10498931. DOI: 10.1002/MPR.329. [Online]. Available: /pmc/articles/PMC3074241/%20/pmc/articles/PMC3074241/?report=abstract%20https://www.ncbi.nlm.nih.gov/pmc/articles/PMC3074241/.

[16] A. Dervishi, “A deep learning backcasting approach to the electrolyte, metabolite, and acid-base parameters that predict risk in icu patients,” PLOS ONE, vol. 15, no. 12, pp. 1–19, Dec. 2020. DOI: 10.1371/journal.pone.0242878.

[17] J. Kasza and R. Wolfe, “Interpretation of commonly used statistical regression models,” Respirology, vol. 19, no. 1, pp. 14–21, 2014. DOI: https://doi.org/10.1111/resp.12221. [Online]. Available: https://onlinelibrary.wiley.com/doi/abs/10.1111/resp.12221.

